# Sputum scarcity and respiratory sample availability among children with presumptive tuberculosis in high burden countries: a systematic review and meta-analysis

**DOI:** 10.1101/2025.11.02.25339327

**Authors:** Mary Gaeddert, Jennifer Habbes, Julian Meister, Ashlyn Beecroft, Amelie von Saint Andre-von Arnim, Beate Kampmann, Mikashmi Kohli, Claudia M. Denkinger, Madhukar Pai, Ankur Gupta-Wright, Florian M. Marx

**Author notes:** Corresponding author: Mary Gaeddert, Im Neuenheimer Feld 324, 69120, Heidelberg, Germany.

## Abstract

**Background:** Tuberculosis (TB) diagnosis typically relies on testing sputum samples, but children often cannot produce sputum. Our review investigated the collection of self-expectorated and induced sputum, and alternative methods of sampling including gastric and nasopharyngeal aspirates, among children evaluated for presumed TB in healthcare facilities.

**Methods:** We searched PubMed, Embase, Cochrane Library, Web of Science, and clinical trials databases from January 2010 to June 2024. Studies not reporting sufficient information on respiratory sampling or not conducted in high TB burden countries were excluded. Summary data was extracted, and the risk of bias was assessed. Sputum scarcity was defined as the proportion of children who could not provide a sample among those attempting. The pooled estimate of sputum scarcity was calculated by random effects meta-analysis. The review protocol was registered with PROSPERO (CRD42023473882).

**Findings:** The search identified 6,751 records and 36 studies were included which enrolled 14,018 children from 14 high burden countries. Respiratory sampling methods varied by age. In children under 5, a pooled estimate of 4% (95% CI: 2-6%, n=5) provided self-expectorated sputum, but collection was not attempted routinely. In studies collecting one or two self-expectorated spot sputum samples in children 5 to 15 years, the pooled estimate of sputum scarcity was 38% (95%CI: 20-55%, n=6). Studies performing sputum induction assisted by nasopharyngeal suctioning in children under 15 had pooled scarcity of 3% (95% CI:0-6%, n=8). For studies performing gastric aspirates, the median proportion without a sample was 0.0% (95% CI: 0·0-0·8%, IQR: 0·0-2·0%, n=23).

**Interpretation:** Collecting respiratory samples in children with presumed TB is complex and age-dependent. Children, especially under 5, are often unable to produce self-expectorated sputum and depend on alternative methods, such as induction or gastric aspirates. TB diagnostics using samples that are easier to collect from children in resource-limited settings are needed.

**Funding:** Gates Foundation (INV-069540)

**Research in Context:** *Evidence before this study:* Current diagnostics for tuberculosis (TB) rely on testing sputum samples, and patients unable to produce sputum at the time of evaluation may have a delayed or missed diagnosis. Children, especially those under 5 years of age, have difficulty producing sputum. There are no currently reliable estimates about how many children being evaluated for presumed TB are unable to produce a sputum sample (sputum scarce).

*Added value of this study:* This systematic review and meta-analysis included data from 36 studies and found that approximately 38% of children between 5 to 15 years of age with presumed TB could not produce one or two sputum samples at the time of evaluation. Alternative methods of respiratory sample collection, such as sputum induction and gastric aspirates, were effective in obtaining samples but are often not available in resource-limited settings.

*Implications of all the available evidence:* Sputum scarcity is common in children being evaluated for TB and alternative methods are necessary to collect respiratory samples. These results support the need to develop non-sputum TB diagnostics for children.

## Introduction

Tuberculosis (TB) represents a serious global health challenge for children. The World Health Organization (WHO) reported that in 2023, an estimated 1·3 million children under 15 years of age developed TB, equivalent to 12% of the estimated total TB burden (1). Among children, 191,000 deaths due to TB occurred in 2023, representing 15% of all TB deaths (1). Over 60% (122,000) of these deaths occurred in children under 5 years of age who often exhibit non-specific signs and symptoms, making diagnosis more challenging (1). Children living with human immunodeficiency virus (CLHIV) are at high risk for HIV-associated TB and have worse treatment outcomes (2), as are children with severe acute malnutrition (SAM) (3).

While TB can be cured, timely diagnosis and treatment among children represents a major challenge (4). Central to this challenge is the difficulty in collecting respiratory samples for microbiologic testing, with primary methods of respiratory sampling varying by age (5). Very young children, typically under 5 years of age, often swallow their respiratory secretions and lack sufficient tussive force to produce sputum samples through self-expectoration. They often require alternative methods of sample collection, such as gastric or nasopharyngeal aspiration or induced sputum with the addition of nasopharyngeal suctioning (6). For children aged 5 to less than 10 years, sputum may be either self-expectorated or require sputum induction using inhaled nebulized saline (7), while older children, from about 10 years old, are usually able to self-expectorate sputum (8).

Gastric aspiration or lavage is used to collect bronchial secretions that have been swallowed. The procedure involves inserting a nasogastric tube and aspirating gastric contents; saline can be inserted down the tube and aspirated to obtain an adequate volume of sample (9). Nasopharyngeal aspirates have also been recommended more recently, which involves nasopharynx aspiration using a mucus trap connected to a suction device, without prior nebulization (10). The string test is another method for gastric sampling in children but is not commonly used in practice. This method requires the child to swallow a weighted gelatin capsule containing an absorbent nylon string while the end is taped to the child’s cheek. The string absorbs secretions in the stomach for several hours, then is carefully withdrawn and processed for testing (11).

However, these alternative methods of sample collection are often not available in many resource-limited settings due to resource and operational challenges (12). In the TB high burden countries, which have the highest incidence of new cases, sample collection is often limited to self-expectorated sputum collected on the spot, i.e. during the same clinical encounter (13). If a child is unable to provide sputum at this time, they may return another time for additional attempts, but are at risk of being lost to follow-up (14). Children who are too young or unable to self-expectorate sputum and do not have access alternative collection methods will usually not have any samples available for microbiologic testing.

Recent policies have prioritized research on the development of non-sputum TB tests that may be easier collect and less invasive in children, including serum, stool, urine, exhaled breath, and oral swabs (15–20). While these samples may have lower sensitivity than respiratory samples, they are potentially easier to collect in resource-limited settings, allowing more children to have a sample for molecular testing, detecting more cases, and resulting in a higher TB diagnostic yield (21).

There is limited knowledge about the extent to which children being evaluated for TB have an adequate respiratory sample collected. The aim of this systematic review and meta-analysis was to investigate sputum scarcity and respiratory sample collection among children being evaluated for presumed TB in healthcare facilities.

## Methods

For this systematic review and meta-analysis, the objective was to describe the proportion of children being evaluated for presumed TB in health facilities who were sputum scarce. We defined sputum scarcity as the proportion of children for whom a sputum sample, either self-expectorated or induced, was attempted but could not be collected. A secondary objective was to investigate the proportion of children for whom a respiratory sample could not be collected using alternative methods (e.g. gastric aspirates, nasopharyngeal aspirates, and the string test).

### Search strategy and selection criteria

Studies were identified through a systematic search of medical databases. The search strategy combined terms for tuberculosis, respiratory samples, and pediatrics (Table S1). PubMed, Embase, Cochrane Library, the Web of Science, Literatura Latino Americana en Ciencias de la Salud (LILACS), clinicaltrials.gov, and International Clinical Trials Registry Platform (ICTRP) were searched and only studies in English were included. We also identified papers from the references of relevant review articles. The search was conducted for all studies from 1 January 2010 to 30 June 2024. In 2010, WHO endorsed the use of Xpert MTB/RIF (Cepheid, USA), a rapid molecular assay for TB diagnosis (12), triggering more intensive research into sputum and non-sputum-based testing for TB. For reasons of feasibility, we did not include studies published before 2010.

Included studies must have enrolled children under 15 years of age with presumptive TB, based on the individual study’s definition. Children without classical TB symptoms but who had risk factors such as HIV or contact with TB cases were also included. The study must have attempted to collect respiratory samples for TB testing, either as part of study procedures or via routine TB diagnostic procedures. Respiratory samples were defined as self-expectorated sputum, induced sputum, gastric aspirates, nasopharyngeal aspirates, and the string test.

Studies investigating non-respiratory samples, such as stool or urine, were included if respiratory samples were also collected. We included randomized clinical trials, cohort studies, and cross-sectional studies, but excluded studies employing active case-finding strategies, prevalence surveys, and household contact studies. Studies enrolling children with only confirmed TB, only extra-pulmonary TB, and children who had already provided respiratory samples prior to screening were excluded. Studies with less than 20 children attempting sample collection and that were not conducted in one of the 30 high TB burden countries were also excluded (13).

Two reviewers independently screened a sample of the titles and abstracts. Given the high degree of concordance between reviewers, only a 5% sample was screened by two reviewers and then one reviewer independently assessed the remaining titles and abstracts, unless they were unsure in which case a second reviewer was consulted.

### Data extraction

A standardized form was used for data extraction. If studies reported using more than one type of respiratory sampling method, data were extracted for each method separately. Study quality was assessed using a bespoke tool adapted from the Quality Assessment of Diagnostic Accuracy Studies (QUADAS-2) (22) and JBI tools (23) (Appendix Table S2). Two reviewers (JH, JM) independently reviewed the full text, extracted data, and completed the quality assessment. Discordance was resolved by decision of a third reviewer (MG). Covidence software (Veritas Health Innovation, Australia) was used for screening, full text review, data extraction, and quality assessment.

### Data analysis and synthesis

Characteristics of the included studies were summarized using quantitative data synthesis. Due to the clinical differences in respiratory sample collection methods, results were analyzed separately by method. For studies collecting self-expectorated and induced sputum, sputum scarcity was defined as the proportion without a sample successfully collected among all children attempting to provide sputum. For studies using alternative methods of sputum collection, such as gastric aspirate, the outcome was measured as the proportion of children unable to provide a clinical specimen among all children undergoing that sampling method. If studies reported results of more than one collection method, proportions were calculated separately for each method.

If there was a high degree of heterogeneity between studies as determined by visual assessment of the forest plots and use of I^2^ statistic, a pooled estimate was not done. Instead, the summary estimate was presented as bootstrapped median with 95% confidence intervals, supplemented by interquartile ranges (IQR).

To estimate the prevalence of sample scarcity and to explain likely sources of heterogeneity, meta-analyses were conducted for sub-groups that had at least four studies. The sub-groups specified *a priori* included CLHIV and SAM. An analysis of self-expectorated sputum by age category (under 5, 5-9, and 10-15) was planned if sufficient data was available. Also, sub-groups focused on the collection of self-expectorated spot sputum samples, as this is the most programmatically relevant (24). We contacted study authors when details of respiratory sampling were not clearly reported.

Data analysis was conducted using Stata (StataCorp. 2024. *Stata Statistical Software: Release 17*. College Station, TX: StataCorp LLC.). We used a random effects meta-analysis with the *meta* command package and the random effects restricted maximum likelihood (REML) method to calculate summary estimates and 95% confidence intervals.

The review protocol and search strategy were registered on in the PROSPERO database (CRD42023473882) and we followed the PRISMA 2020 guidelines for reporting systematic reviews and meta-analyses (25).

## Results

The literature search resulted in 6,751 articles after de-duplication. After title and abstract screening, 171 were eligible for full-text review. Of these, 135 were excluded, most commonly for not being conducted in a high-burden country (n=41) or not adequately reporting details of respiratory sample collection (n=39), leaving 36 studies in the systematic review (Figure 1).

**Figure 1.**
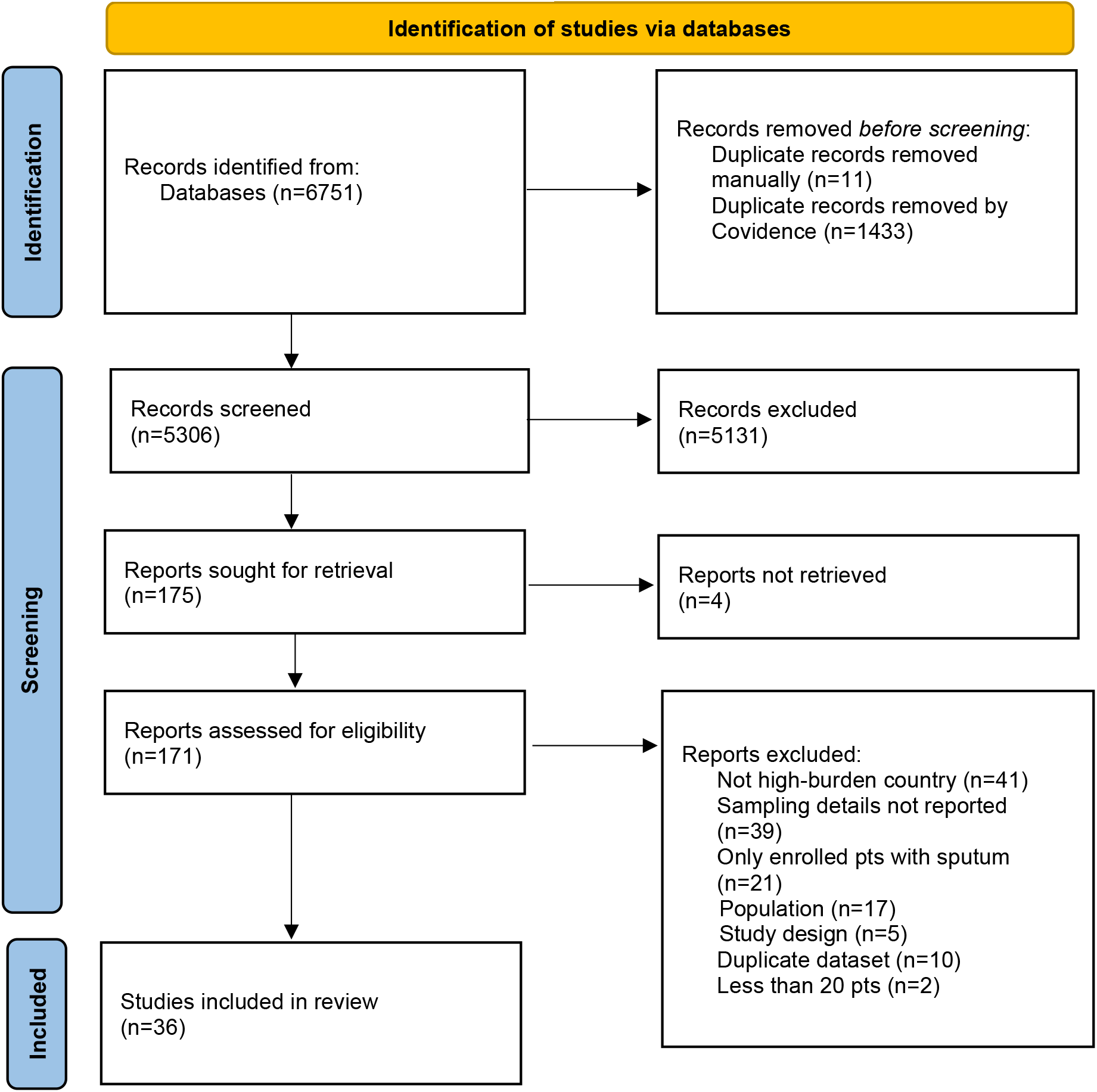
PRISMA flow diagram.

The included studies enrolled a total of 14,018 children from 14 different countries. The majority of studies were cross-sectional in design (n=33, 91·7%), evaluated the accuracy of TB diagnostic tests (n=23, 63·9%) and were conducted at tertiary level hospitals (n=28, 77·8%) (Table 1, details in Table S3). Studies enrolled children from outpatient (n=4, 11·1%), inpatient (n=10, 27·8%), or mixed inpatient and outpatient settings (n=10, 27·8%), and the remaining studies did not report the setting (n=12, 33·3%). Studies used multiple criteria to identify children with presumptive TB, including symptoms (n=33, 91·7%), contact with a TB case (n=20, 55·6%), abnormal chest X-ray findings (n=17, 47·2%) and being in a high-risk group such as CLHIV (n=5, 13·9%). A median 19·0% (IQR: 13·0-31·0%, n=25 studies) were reported as CLHIV, and a median 35·1% (IQR: 16·0-49·0%, n=20 studies) of children were reported to have SAM. Two studies enrolled only CLHIV and one study enrolled only children with SAM. The median age of children was 32 months (IQR: 24-48 months, n=25 studies), and a median 67·4% (IQR: 53·7-89·3%, n=20 studies) of children were age 5 and under. Reporting of age was heterogeneous and 11 (30·1%) studies did not report median age.

**Table 1.** Summary of included studies.

| **Characteristic** | **N (%)** |
| --- | --- |
| Study Design |  |
| Cross-sectional | 33 (91.7) |
| Cohort | 2 (5.6) |
| Randomized controlled trial | 1 (2.8) |
| Diagnostic accuracy study | 23 (63.9) |
| Study enrollment by WHO region |  |
| African | 22 (61.1) |
| South-East Asian | 10 (27.8) |
| Western Pacific | 2 (5.6) |
| Americas | 1 (2.8) |
| Eastern Mediterranean | 1 (2.8) |
| Clinical setting |  |
| Mixed out/inpatient | 10 (27.8) |
| Inpatient | 10 (27.8) |
| Outpatient | 4 (11.1) |
| Not reported | 12 (33.3) |
| Level of care |  |
| Tertiary | 28 (77.8) |
| Mixed levels | 4 (11.1) |
| Primary | 2 (5.6) |
| Not reported | 2 (5.6) |
| Criteria for presumptive TB* |  |
| TB Symptoms | 33 (91.7) |
| Chest X-ray | 17 (47.2) |
| TST-positive | 2 (5.6) |
| High-risk group | 27 (75.0) |
| High-risk groups* |  |
| Contact with TB case | 20 (55.6) |
| Malnutrition | 4 (11.1) |
| CLHIV | 5 (13.9) |
| Respiratory sample collection method* |  |
| Gastric aspirate | 19 (52.8) |
| Induced sputum | 19 (52.8) |
| Self-expectorated sputum | 15 (41.7) |
| Nasopharyngeal aspirate | 4 (11.1) |
| String test | 3 (8.3) |
| Number samples attempted to collect |  |
| 1 | 14 (38.9) |
| 1-2 | 13 (36.1) |
| >2 | 3 (8.3) |
| 2 of each sample type | 5 (13.9) |
| Not reported | 1 (2.8) |
| Duration of sample collection |  |
| Spot | 10 (27.8) |
| 1-2 days | 10 (27.8) |
| >2 days | 2 (5.6) |
| Not reported | 14 (38.9) |
| **Study-level population characteristics** | **Median [IQR]** |
| TB prevalence (n=36 studies) | 11.1% [6.5-18.8] |
| HIV prevalence (n=25 studies) | 19.0% [13.0-31.0] |
| CLHIV, on ART (n=9 studies) | 57.0% [25.0-84.2] |
| History of TB disease (n=9 studies) | 7.2% [2.5-9.5] |
| Severe acute malnutrition (n=20 studies) | 35.1% [16.0-49.0] |
| Age, months (n=25 studies) | 32.0 [24.0-48.0] |

\*categories are not mutually exclusive

Most studies (n=22, 61·1%) collected more than one type of respiratory sample, and the sampling strategy was based on age. Children under 5 years frequently had gastric aspirates or induced sputum and expectorated sputum was only attempted routinely in children over 5. If children were unable to self-expectorate, then gastric aspirates, induced sputum, or nasopharyngeal aspirates were collected. However, the details of sample collection by age were only noted in the text for 8 (22·2%) studies and it was necessary to contact authors to obtain additional information.

Overall, 19 studies (52·8%) each collected gastric aspirates and induced sputum, either as the primary method or in sequence. Of the studies collecting induced sputum, 11 (57·9%) reported performing nasopharyngeal suctioning if the child was unable to produce a sample by coughing after nebulization. Fifteen (41·7%) studies attempted to collect self-expectorated sputum. Nasopharyngeal aspirates (n=4, 11·1%) and the string test (n=3, 8·3%) were less common. Most studies attempted to collect one or two samples (n=27, 75·0%) either on the spot (e.g. in one clinical encounter) (n=10, 27·8%) or over the course of one to two days of clinical visits (n=10, 27·8%). However, 14 (38·9%) studies did not report the time over which samples were collected.

The risk of bias assessment scored 35 (97·2%) studies as low risk of bias on the patient selection domain (Figure 2). These studies enrolled a consecutive or random sample of participants, did not use a case-control design, and avoided inappropriate exclusions. For the applicability domain, 11 (30·6%) scored as a high risk of bias were due to not reporting details on how respiratory samples were collected.

**Figure 2.**
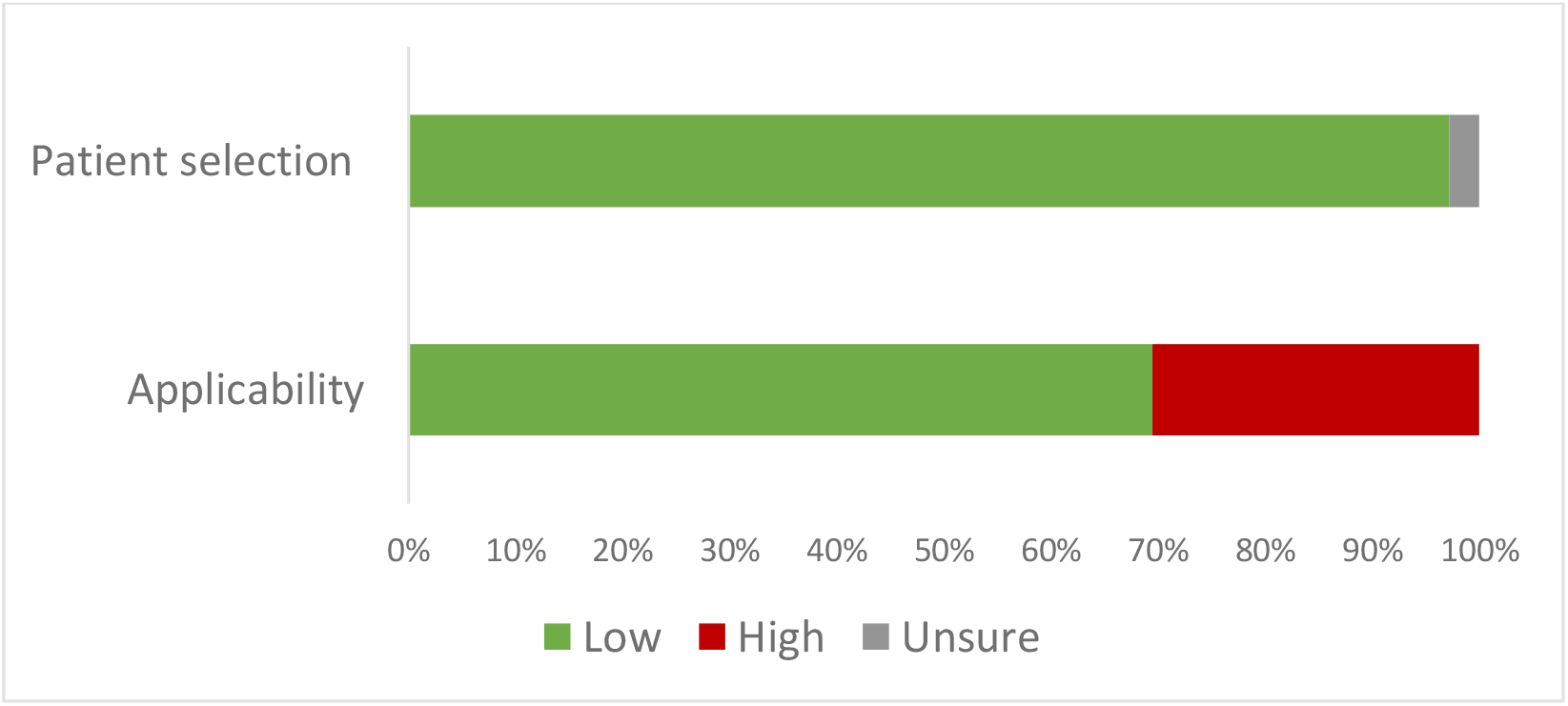
Summary of risk of bias assessment.

Sub-group meta-analyses were performed for self-expectorated and induced sputum. Limited data was reported on sample collection by the pre-determined age categories, so self-expectorated sputum was analyzed for children under 5 and 5 to 15 years of age separately, and induced sputum was analyzed for all children under 15. In studies which reported collecting self-expectorated sputum from children under 5, the pooled estimate of 4% (95% CI: 2-6%, n=5) were able to provide samples (Figure S1). However, self-expectorated sputum was not attempted routinely in this age group and only done based on physician discretion or the presence of a productive cough.

In studies collecting self-expectorated sputum from children 5 to 15 years of age, the pooled estimate of sputum scarcity for collection of any sputum sample was 33% (95% CI: 22-44%, n=11) (Figure S2). When restricted to studies collecting one or two self-expectorated spot sputum samples in children 5-15 years, the pooled scarcity estimate was 38% (95%CI: 20-55%, n=6) (Figure S3). In studies that reported HIV status, the pooled estimate of self-expectorated sputum scarcity for children 5-15 years was 31% (95%CI: 20-42%, n=7) for all sample numbers and times (Figure S4). The median proportion of CLHIV in these studies was 30·5% (IQR: 18·0-32·9%). And in studies that reported enrolling children with severe acute malnutrition, the pooled estimate of sputum scarcity for collection of one or two self-expectorated spot samples in children 5-15 was 31% (95% CI: 17-45%, n=5) (Figure S5). The median proportion of SAM in these studies was 58·6% (IQR: 19·0-84·0%).

For studies performing sputum induction in children under 15 years, the pooled estimate of scarcity was 3% (95% CI: 1-5%, n=14) (Figure S6) and was similar for the studies that reported using suctioning if the child was unable to cough (pooled estimate 3%, 95% CI: 0-6%, n=8) (Figure S7). In two studies that reported results, 31·9% of samples were collected by coughing, and the remaining 68·1% were collected by suctioning (7, 26). The subgroup results are summarized in Figure 3.

**Figure 3.**
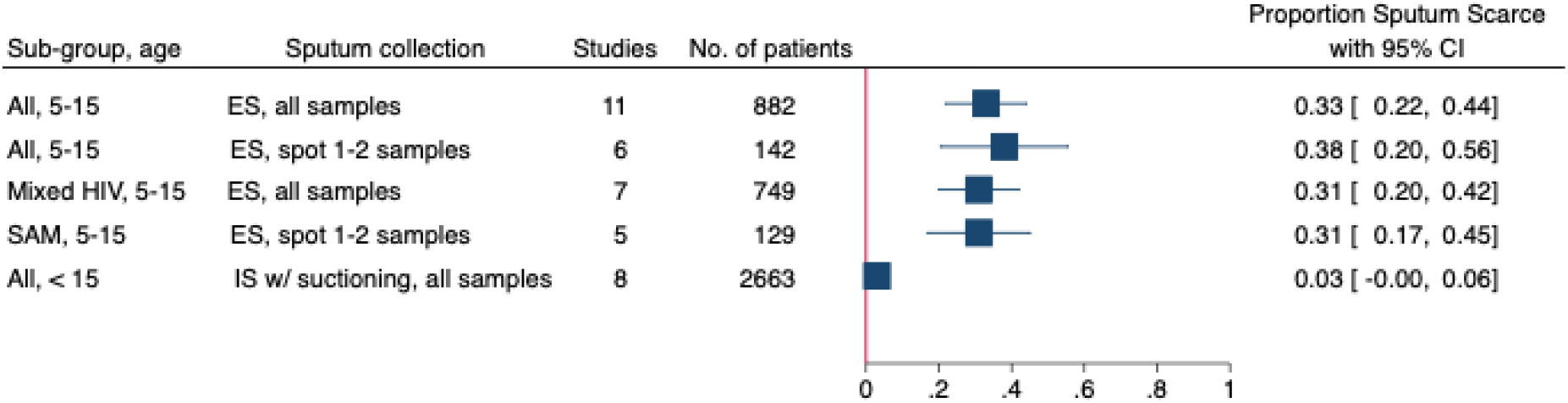
Meta forest plot for proportion of sputum scarcity in sub-groups. Legend: ES=self-expectorated sputum, IS=induced sputum, SAM=severe acute malnutrition

Studies using alternative sampling methods such as gastric aspirates and nasopharyngeal aspirates were almost always successful in collecting a sample. The median proportion without a sample available in studies performing gastric aspirates was 0·0% (95% CI: 0·0-0·8%, IQR: 0·0-2·0%, n=23) (Table 2). In the three studies reporting results of nasopharyngeal aspirates, the median proportion without a sample available was 3·6% (95% CI: 3·4-3·9%, IQR: 3·5-3·8%). Additionally, for three studies evaluating the string test, the median proportion without a sample was 16·1% (95% CI: 3·6-28·5%, IQR: 3·1-21·2%).

**Table 2.** Proportion of children without sample available, by sampling method.

| Respiratory sampling method | Number of studies | Number of patients | Proportion without sample, Median (95% CI) | IQR |
| --- | --- | --- | --- | --- |
| Gastric aspirate | 23 | 9,679 | 0.00 (0.00, 0.08) | [0.00-0.02] |
| Nasopharyngeal aspirate | 3 | 306 | 0.04 (0.03, 0.04) | [0.03-0.04] |
| String test | 3 | 464 | 0.16 (0.04, 0.28) | [0.03-0.21] |

## Discussion

This systematic review and meta-analysis reports the first estimates on sputum scarcity in children. Our main finding is that approximately 30% of children between 5 to 15 years of age being evaluated for presumed TB in high burden countries are unable to produce at least one self-expectorated sputum sample at the time of evaluation. Children younger than 5 years depend on alternative methods, such as gastric aspirates, as only a small number can self-expectorate. When sputum induction was performed, only 3% of children could not produce a sample, although this was often assisted with suctioning if the child was unable to cough. Studies collecting gastric and nasopharyngeal aspirates were successful in obtaining a sample in almost all children.

While alternative sampling methods are effective, the resources required limit their use mainly to tertiary level hospitals, where 78% of these studies were conducted. All sampling methods require trained staff and expertise working with young children. Gastric aspirates should be performed after overnight fasting, usually require hospital admission with trained personnel and repeat sampling, and can be uncomfortable for the child (9). Sputum induction requires nebulizer equipment and airborne infection control due to the generation of aerosols (8). Sputum induction is generally well-tolerated but can be difficult for some children (7) and nasopharyngeal suctioning may be stressful. The string test does not require significant resources, but was noted to not be well-tolerated, as children can have difficulty swallowing the gelatin capsule and reported discomfort during insertion and removal of the string (11).

Even when a respiratory sample can be obtained, the sensitivity of molecular testing is reduced due to the paucibacillary nature of childhood TB disease (27). A review of Xpert diagnostic accuracy for childhood TB found the pooled sensitivity of Xpert Ultra compared to culture was 73% in children without HIV and 66% in CLHIV (28). Another review showed the detection yields of alternative sampling methods in children were low and varied by sampling method. The yield of Xpert for the first sample was 2-17% for induced sputum, 5-51% for gastric aspirate, and 8% for nasopharyngeal aspirate. Yield increased when collecting a second sample or combining methods of sample collection (29).

If no sample is available for microbiological testing, or if the sample is negative, diagnosis will rely on clinical signs and symptoms, which has limited accuracy and cannot provide information on drug resistance (30). Also, many clinicians outside of referral centers may be reluctant to treat children without bacteriological confirmation due to lack of expertise (31). In recognition of these challenges, stool and urine samples are now recommended for molecular testing in children (4). The sensitivity of stool testing with Xpert Ultra is 56% (28) and the sensitivity of Alere LAM for CLHIV is 47% (17).

While non-sputum samples may have lower sensitivity than sputum, the ability to collect samples from more children may increase diagnostic yield. For example, the implementation of stool testing in Zambia expanded access to bacteriological testing for children under 5 and increased the bacteriological confirmation rate by 53% (32). Our estimates of how many children cannot provide self-expectorated sputum and would benefit from non-sputum samples will enable more accurate modelling of the potential impact of new diagnostics for childhood TB, especially in low-resource settings without access to alternative sample collection methods.

Strengths of this review were the comprehensive literature search and screening methods, and the large number of studies included. All papers were from studies conducted in high TB burden countries and had a low risk of bias for participant selection. However, incomplete reporting of key variables limited our analysis. Results of sample collection were often not reported separately when multiple techniques were used, so it was necessary to contact authors to obtain additional data, especially for self-expectorated sputum. Different age categories and definitions of malnutrition were used, and sample quality was not reported in any studies. As most studies were conducted in tertiary care centers, the generalizability of the results to lower levels of care and outpatient settings is limited. Our results may be an underestimate of the true burden of sputum scarcity in peripheral settings.

Further development of child-friendly samples and collection methods, such as stool and tongue swabs, are needed to improve diagnostic capacity for childhood TB in low-resource settings. Studies in children should consistently report the respiratory sampling methods used and the results of sample collection attempts.

While is widely held that children have difficulty producing sputum, this review quantifies the extent of sputum scarcity and reliance on alternative sample collection methods. These findings provide valuable insight into a high-risk group that would greatly benefit from non-sputum tests, supporting further development and implementation of child-friendly sampling strategies.

## Data Availability

All data produced in the present work are contained in the manuscript.

## Declarations

### Contributors

AGW, FM, MP, and CMD developed the idea and initiated the article. MG, FM, and AGW, AvStA, and BK drafted study design and protocol with input from CMD. The search was designed and conducted by MG. Screening, data extraction and risk of bias assessment was done by MG, JH, JM. MG conducted the data analysis. AGW, FM, MG, AB, MK, MP, and CMD interpreted results. MG wrote the first draft of the manuscript. All authors contributed to the manuscript by edits and providing critical feedback. All authors had full access to the data and had final responsibility for submission.

### Data sharing

This manuscript is based on secondary data which is published and publicly available. Data extracted for the analysis is presented in the results section and Supplementary Appendix. The code used for the analysis is available upon request.

## Acknowledgements

We would like to thank all the authors who provided additional information and results on their studies. We would also like to thank Stefan Weber for reviewing the manuscript.

This work was supported, in whole or in part, by the Gates Foundation [INV-069540]. The conclusions and opinions expressed in this work are those of the author(s) alone and shall not be attributed to the Foundation. Under the grant conditions of the Foundation, a Creative Commons Attribution 4.0 License has already been assigned to the Author Accepted Manuscript version that might arise from this submission. Please note works submitted as a preprint have not undergone a peer review process.

MG and AB are supported by the Gates Foundation (award number INV-069540). CMD and AGW are funded by the R2D2 TB Network (National Institute of Allergy and Infectious Diseases of the US National Institutes of Health under award number U01AI152087). AGW is supported by the UK National Institute for Health and Care Research (NIHR305136) and Academy of Medical Sciences (SGCL025\1043).

