## Supplemental materials for "Sputum scarcity and respiratory sample availability among children with presumptive tuberculosis in high burden countries: a systematic review and meta-analysis"

**Supplemental Material**

### Table S1. Search Strategy

| **PubMed** | |
|---|---|
| #1 | "Mycobacterium tuberculosis"[MeSH] OR "Tuberculosis"[MeSH] OR Tuberculo*[tiab] OR TB[tiab] |
| #2 | "sputum"[MeSH] OR sputum[tiab] OR "respiratory aspiration"[MeSH] OR "gastric aspirat*"[tiab] OR "gastric lavag*"[tiab] OR "nasopharyngeal aspirat*"[tiab] OR "string test*"[tiab] OR sputa[tiab] OR expectorat*[tiab] OR phlegm*[tiab] |
| #3 | "Urine"[Mesh] OR "Feces"[Mesh] OR "Blood"[Mesh] OR "Serum"[Mesh] OR urin*[tiab] OR stool[tiab] OR fece*[tiab] OR blood[tiab] OR serum*[tiab] OR swab*[tiab] |
| #4 | #2 OR #3 |
| #5 | "child"[Mesh] OR "pediatrics"[Mesh] OR childhood*[tiab] OR child*[tiab] OR pediatric*[tiab] OR paediatric*[tiab] OR infan*[tiab] |
| #7 | #1 AND #4 AND #5 |
| #8 | "2010/01/01"[PDat] : "2024/6/30"[PDat] |

| **Embase** | |
|---|---|
| #1 | 'Mycobacterium tuberculosis'/exp OR 'tuberculosis'/exp OR Tuberculo*:ti,ab,kw OR TB:ti,ab,kw |
| #2 | 'sputum'/exp OR 'sputum examination'/exp OR sputum*:ti,ab,kw OR aspirat*:ti,ab,kw |
| #3 | 'urine'/exp OR 'feces'/exp OR 'blood'/exp OR 'serum'/exp OR 'feces analysis'/exp OR 'urinalysis'/exp OR urin*:ti,ab,kw OR stool:ti,ab,kw OR fece*:ti,ab,kw OR blood:ti,ab,kw OR serum*:ti,ab,kw OR swab*:ti,ab,kw |
| #4 | #2 OR #3 |
| #5 | child*:ti,ab,kw OR pediatric*:ti,ab,kw OR paediatric*:ti,ab,kw OR infan*:ti,ab,kw |
| #6 | #1 AND #4 AND #5 |
| | Publication date filter: 2010-2024 |
| | Filter: Embase only |
| | Filter: all categories under 18 |
| | Filter: humans, only English |

**Table S2. Template for Risk of Bias assessment**

| **Domain**<br>**Signaling Question** | **Accepted values and answers** |
|---|---|
| **Domain 1: Patient Selection**<br>Could the selection of patients have introduced bias? | |
| 1. Was a consecutive or random sample of patients enrolled? | Random → yes<br>Consecutive → yes<br>Convenience → no<br>NR → unsure |
| 2. Was a case-control design avoided? | Cohort → yes<br>Cross-sectional → yes<br>Case-control → no<br>NR → unsure |
| 3. Did the study avoid inappropriate exclusions? (e.g. previous TB, HIV, unable to provide an ERS) | Yes → yes<br>No → no<br>NR → unsure |
| Scoring:<br>Yes on ≥ 2 questions → Low<br>No on ≥ 2 questions → High<br>Unsure on ≥ 2 questions → Unsure | |
| **Domain 2: Assessment of the Outcome**<br>Is there a concern that the included participants do not match the review question? | |
| 1. Were methods of ERS collection described in sufficient detail? | Yes → yes<br>No → no<br>NR → unsure |
| 2. Were the results of ERS collection available for all, or nearly all, participants? | Yes → yes<br>No → no<br>NR → unsure |
| 3. Was information on sputum quality (e.g. salivary samples) reported? | Yes → yes<br>No → no<br>NR → unsure |
| Scoring:<br>Yes on ≥ 2 questions → Low<br>No on ≥ 2 questions → High<br>Unsure on ≥ 2 questions → Unsure | |

### Table S3. Characteristics of included studies

| Study ID | Design; DTA | Country (-ies) | Clinical setting | Healthcare level | N pts | TB prevalence (%) | HIV prevalence (%) | Previous TB (%) | SAM prevalence (%) | Collection method | Number samples | Collect time | Proportion scarcity/ without sample | RoB |
|---|---|---|---|---|---|---|---|---|---|---|---|---|---|---|
| Ainan 2021 (1) | CSS; Yes | Tanzania | Mixed | Mixed | 253 | 3.6 | 6.5 | NR | NR | ES, GA | 2 | NR | GA: 0.04 | Low, High |
| Bates 2013 (2) | CSS; Yes | Zambia | Inpatient | Tertiary | 1037 | 6.2 | 30.5 | NR | NR | ES, GA | NR | NR | ES: 0.52<br>GA: 0.00<br>ES<5: 15/663 with sample | Low, Low |
| Cardoso 2022 (3) | CSS; No | Brazil | Mixed | Tertiary | 33 | 19.0 | NR | NR | NR | ST | 1 | Spot | ST: 0.21 | Low, Low |
| Chibolela 2023 (4) | CSS; No | Zambia | NR | Tertiary | 116 | 11.4 | 23.7 | NR | 35.1 | GA | 1 | NR | GA: 0.00 | Low, Low |
| Cox 2022 (5) | CSS; Yes | South Africa | NR | Tertiary | 328 | 30.9 | 19.6 | 7.6 | 29.9 | IS w/suction | 2 | 1-2 days | IS: 0.02 | Low, High |
| Dayal 2021 (6) | CSS; Yes | India | NR | Tertiary | 114 | 51.7 | NR | NR | 10.5 | IS, GA | 1 | Spot | GA: 0.00 | Low, High |
| Dubale 2022 (7) | CSS; Yes | Ethiopia | NR | Tertiary | 152 | 6.7 | NR | NR | 35.5 | ES, GA | 1 | NR | GA: 0.00 | Low, High |
| Ebonyi 2020 (8) | CSS; Yes | Nigeria | Outpatient | Tertiary | 103 | 45.6 | NR | NR | 68.7 | ES, GA | 1 | Spot | ES: 0.29<br>GA: 0.00<br>ES<5: 4/58 with sample | Low, High |
| Hanrahan 2019 (9) | Cohort; No | South Africa | Outpatient | Primary | 119 | 3.0 | 18.0 | 4.0 | 16.0 | ES, NPA, IS, GA | 1 ES, 2 NPA, 1 IS, 1 GA | Spot | ES: 0.41<br>IS: 0.02<br>GA: 0.23<br>NPA: 0.04<br>ES<5: 4/102 with sample | Low, Low |
| Hasan 2017 (10) | CSS; Yes | Pakistan | NR | Tertiary | 50 | 18.0 | NR | NR | NR | ES, GA | 1 | NR | ES: 0.69<br>GA: 0.00 | Low, Low |
| Jaganath 2021 (11) | CSS; Yes | Uganda | Mixed | Tertiary | 217 | 10.8 | 13.0 | 1.4 | NR | ES, IS, NPA, GA | 2 | NR | ES: 0.54 | Low, High |
| Kabir 2018 (12) | CSS; Yes | Bangladesh | Inpatient | Tertiary | 102 | 15.7 | NR | NR | 86.2 | ES, GA | 1 | Spot | ES: 0.64<br>GA: 0.00 | Low, Low |
| Kabir 2021 (13) | CSS; Yes | Bangladesh | Inpatient | Tertiary | 447 | 16.1 | NR | 2.5 | 41.8 | IS w/ suction | 1 | NR | IS: 0.00 | Low, Low |
| KasaTom 2018 (14) | CSS; Yes | Papua New Guinea | Inpatient | Tertiary | 100 | 28.0 | 18.3 | NR | 48.4 | ES, GA | 2 ES, 1 GA | Spot | ES: 0.15<br>GA: 0.05<br>ES<5: 3/60 with sample | Low, Low |

| | | | | | | | | | | | | | | |
|---|---|---|---|---|---|---|---|---|---|---|---|---|---|---|
| Khambati 2024 (15) | CSS; No | Kenya | Mixed | Mixed | 300 | 10.5 | 24.3 | NR | NR | GA, ST | 2 each | 3 days | GA: 0.02<br>ST: 0.03 | Low, Low |
| Kroidl 2015 (16) | CSS; Yes | Tanzania | Outpatient | Tertiary | 180 | 13.6 | 51.0 | NR | NR | IS | 3 | NR | IS: 0.01 | Low, Low |
| LaCourse 2018 (17) | RCT; Yes | Kenya | Inpatient | Tertiary | 181 | 7.9 | 100 | NR | 48.1 | IS, GA | 2 | 1-2 days | GA: 0.04 | Low, Low |
| Menon 2011 (18) | CSS; No | India | NR | Tertiary | 52 | 36.5 | 0 | NR | NR | GA | Up to 3 | NR | GA: 0.00 | Low, Low |
| Moore 2011 (19) | CSS; No | South Africa | NR | Primary | 270 | 10.7 | 18.0 | NR | NR | IS w/suction | 2 | NR | IS: 0.01* | Low, Low |
| Moore 2017 (20) | CSS; No | South Africa | Inpatient | Tertiary | 920 | 2.9 | 12.5 | NR | 15.9 | IS, GA | 2 | 1-2 days | IS: 0.09<br>GA: 0.02 | Low, Low |
| Mutabazi 2020 (21) | CSS; No | Tanzania | Mixed | Mixed | 263 | 5.2 | 100 | 9.5 | 4.4 | ES, GA | 2 ES, 1 GA | 1-2 days | ES: 0.15<br>GA: 0.07 | Low, Low |
| Myo 2018 (22) | CSS; Yes | Myanmar | NR | Tertiary | 231 | 16.5 | 19.0 | 13.0 | NR | GA | 1 | NR | GA: 0.00 | Low, Low |
| Nansumba 2016 (23) | CSS; No | Uganda | Mixed | Tertiary | 137 | 10.2 | 31.0 | NR | 3.6 | ST, IS w/ suction | 2 each | 2 days | IS: 0.08<br>ST: 0.16 | Low, Low |
| Nicol 2019 (24) | CSS; Yes | South Africa | NR | Tertiary | 165 | 24.2 | 10.9 | NR | NR | IS w/suction | 2 | NR | IS: 0.00 | Low, Low |
| Orikiriza 2018 (25) | Cohort; Yes | Uganda | Mixed | Tertiary | 385 | 4.3 | 31.2 | 2.0 | 19.0 | ES, IS w/suction | 2 | 2 days | ES: 0.36<br>IS: 0.11 | Low, High |
| Orikiriza 2022 (26) | CSS; Yes | Uganda | Inpatient | Tertiary | 213 | 5.5 | 32.9 | NR | 84.0 | ES, IS w/ suction, NPA, GA | 2 | 2 days | ES: 0.22<br>NPA: 0.03<br>GA: 0.03<br>ES<5: 5/39 with sample | Low, High |
| Pang 2014 (27) | CSS; Yes | China | NR | Tertiary | 211 | 8.1 | NR | NR | NR | GA | 1 | Spot | GA: 0.00 | Low, Low |
| Planting 2014 (28) | CSS; No | South Africa | Inpatient | Tertiary | 843 | 18.6 | 23.8 | 10.4 | 15.6 | IS w/ suction | 2 | 1-2 days | IS: 0.02* | Low, Low |
| Sabi 2016 (29) | CSS; No | Tanzania | Mixed | Tertiary | 192 | 5.2 | 15.1 | NR | 49.5 | IS w/ suction | 1 | NR | IS: 0.03 | Low, Low |
| Sekadde 2013 (30) | CSS; Yes | Uganda | Mixed | Tertiary | 255 | 14.0 | 41.6 | 7.2 | 27.2 | IS | 1 | Spot | IS: 0.02 | Low, Low |
| Singh 2021 (31) | CSS; Yes | India | Outpatient | Tertiary | 356 | 27.4 | NR | NR | NR | IS, GA | 2 each | 2 days | IS: 0.06<br>GA: 0.00 | Low, High |
| Singh 2023 (32) | CSS; No | India | Inpatient | NR | 4356 | 2.0 | 0 | NR | 100 | GA | 1 | NR | GA: 0.02 | Low, Low |
| Sreedeep 2020 (33) | CSS; Yes | India | NR | Tertiary | 55 | 13.3 | 11.7 | NR | 35.0 | ES, GA | 2 | 2 days | GA: 0.00 | Low, Low |
| Tiwari 2015 (34) | CSS; Yes | India | NR | NR | 100 | 36.0 | NR | NR | NR | ES, GA | 2 ES, 3 GA | 2-3 days | ES: 0.08<br>GA: 0.00 | Low, High |
| Yalamanchi 2023 (35) | CSS; No | India | Inpatient | Tertiary | 255 | 10.2 | NR | NR | NR | IS, GA | 2 each | 2 days | IS: 0.02<br>GA: 0.00 | Unsure High |

| | | | | | | | | | | | | | | |
|---|---|---|---|---|---|---|---|---|---|---|---|---|---|---|
| Yenew 2024 (36) | CSS; Yes | Ethiopia | Mixed | Mixed | 896 | 7.6 | 13.2 | NR | NR | ES, GA | 1 | Spot | GA: 0.00 | Low, Low |
| Studies reporting results of suctioning if unable to cough after nebulization:<br>Moore 2011: Of the 496 IS procedures, 296 (60%) samples were obtained by coughing and 200 (40%) by suctioning.<br>Planting 2014: Of the 1257 IS procedures, 264 (21.0%) were obtained by coughing and 993 (79.0%) by suctioning. | | | | | | | | | | | | | | |

**Legend**: (abbreviations)
Design: CSS=cross-sectional study; RCT=randomized controlled trial
DTA=diagnostic test accuracy (yes, no)
Clinical setting: inpatient, outpatient, mixed inpatient and outpatient, not reported (NR)
Healthcare level: Primary, Secondary, Tertiary, mixed levels
N pts: number of participants attempting respiratory sample collection
SAM = severe acute malnutrition
Collection method: self-expectorated sputum (ES), induced sputum (IS), with suctioning if child unable to cough (w/suction), gastric aspirate (GA), nasopharyngeal aspirate (NPA), self-expectorated sputum in children <5 years (ES<5)
Number of samples attempted: 1-2, more than 2, not reported (NR)
Time of sample collection: spot, 1-2 days, 3 days or more, not reported (NR)
Risk of Bias (RoB): Patient selection, Applicability

**Figure S1. Meta-analysis of proportion children under 5 years able to provide self-expectorated sputum**

| Study | Method | No. of samples | Collect time | No. of patients | Proportion with sputum with 95% CI |
|---|---|---|---|---|---|
| Bates 2013 | ES | . | . | 663 | 0.02 [ 0.01, 0.03] |
| Hanrahan 2019 | ES | 1 | Spot | 102 | 0.04 [ 0.00, 0.08] |
| KasaTom 2018 | ES | 2 | Spot | 60 | 0.05 [ -0.01, 0.11] |
| Ebonyi 2020 | ES | 1 | Spot | 58 | 0.07 [ 0.00, 0.13] |
| Orikiriza 2022 | ES | 2 | Spot | 39 | 0.13 [ 0.02, 0.23] |
| **Overall** | | | | | 0.04 [ 0.02, 0.06] |

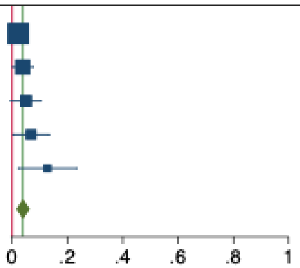


**Figure S2. Meta-analysis of proportion sputum scarcity for collection of self-expectorated sputum in children 5-15 years**

| Study | Method | No. of samples | Collect time | No. of patients | Proportion Sputum Scarce with 95% CI |
|---|---|---|---|---|---|
| Tiwari 2015 | ES | 2 | >2 days | 64 | 0.08 [ 0.01, 0.15] |
| KasaTom 2018 | ES | 2 | Spot | 33 | 0.15 [ 0.03, 0.27] |
| Mutabazi 2020 | ES | 2 | 1-2 days | 191 | 0.15 [ 0.10, 0.20] |
| Orikiriza 2022 | ES | 2 | Spot | 23 | 0.22 [ 0.05, 0.39] |
| Ebonyi 2020 | ES | 1 | Spot | 45 | 0.29 [ 0.16, 0.42] |
| Jaganath 2021 | ES | 2 | . | 59 | 0.36 [ 0.23, 0.48] |
| Orikiriza 2018 | ES | . | 1-2 days | 164 | 0.36 [ 0.29, 0.43] |
| Hanrahan 2019 | ES | 1 | Spot | 17 | 0.41 [ 0.18, 0.65] |
| Bates 2013 | ES | . | . | 262 | 0.52 [ 0.46, 0.58] |
| Kabir 2018 | ES | 1 | Spot | 11 | 0.64 [ 0.35, 0.92] |
| Hasan 2017 | ES | 1 | . | 13 | 0.69 [ 0.44, 0.94] |
| **Overall** | | | | | 0.33 [ 0.22, 0.44] |

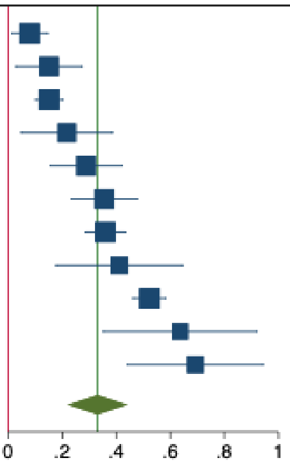


**Figure S3. Meta-analysis of proportion sputum scarcity for collection of 1-2 self-expectorated spot samples in children 5-15 years**

| Study | Method | No. of samples | Collect time | No. of patients | Proportion Sputum Scarce with 95% CI |
|---|---|---|---|---|---|
| KasaTom 2018 | ES | 2 | Spot | 33 | 0.15 [ 0.03, 0.27] |
| Orikiriza 2022 | ES | 2 | Spot | 23 | 0.22 [ 0.05, 0.39] |
| Ebonyi 2020 | ES | 1 | Spot | 45 | 0.29 [ 0.16, 0.42] |
| Hanrahan 2019 | ES | 1 | Spot | 17 | 0.41 [ 0.18, 0.65] |
| Kabir 2018 | ES | 1 | Spot | 11 | 0.64 [ 0.35, 0.92] |
| Hasan 2017 | ES | 1 | . | 13 | 0.69 [ 0.44, 0.94] |
| **Overall** | | | | | 0.38 [ 0.20, 0.55] |

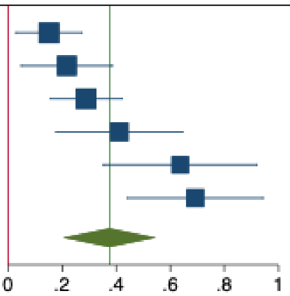

**Figure S4. Meta-analysis of proportion sputum scarcity in studies with mixed HIV population for collection of self-expectorated sputum in children 5-15 years**

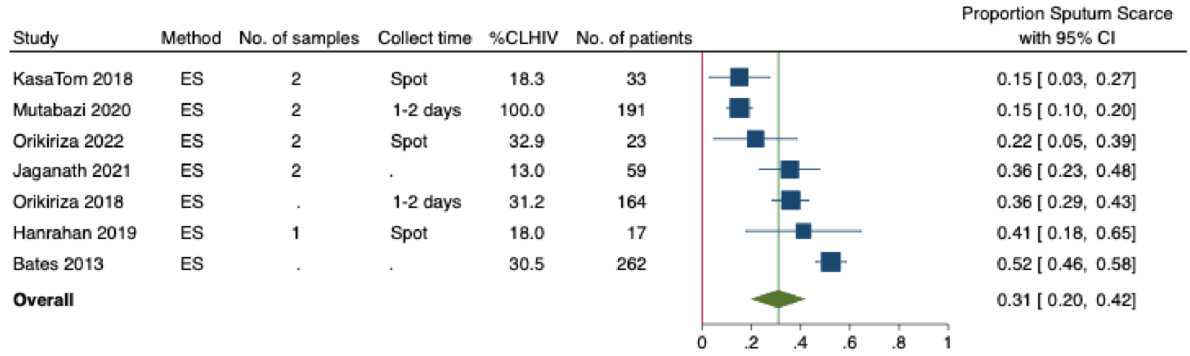


**Figure S5. Meta-analysis of proportion sputum scarcity in studies with mixed SAM population for collection of self-expectorated sputum in children 5-15 years**

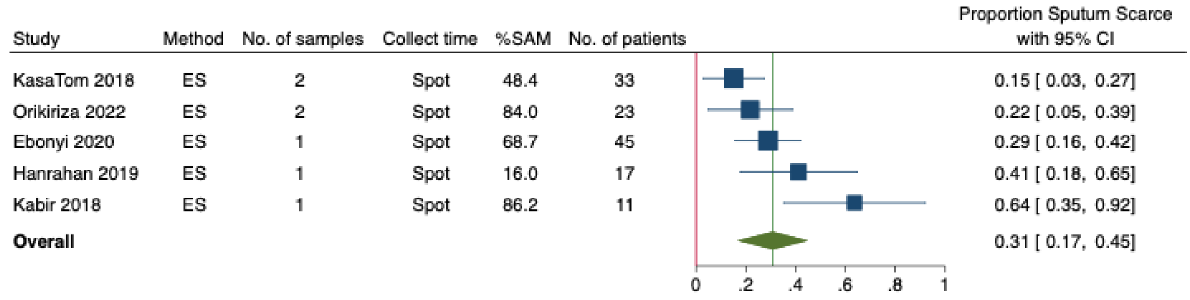


**Figure S6. Meta-analysis of proportion sputum scarcity for collection of induced sputum, children<15 years**

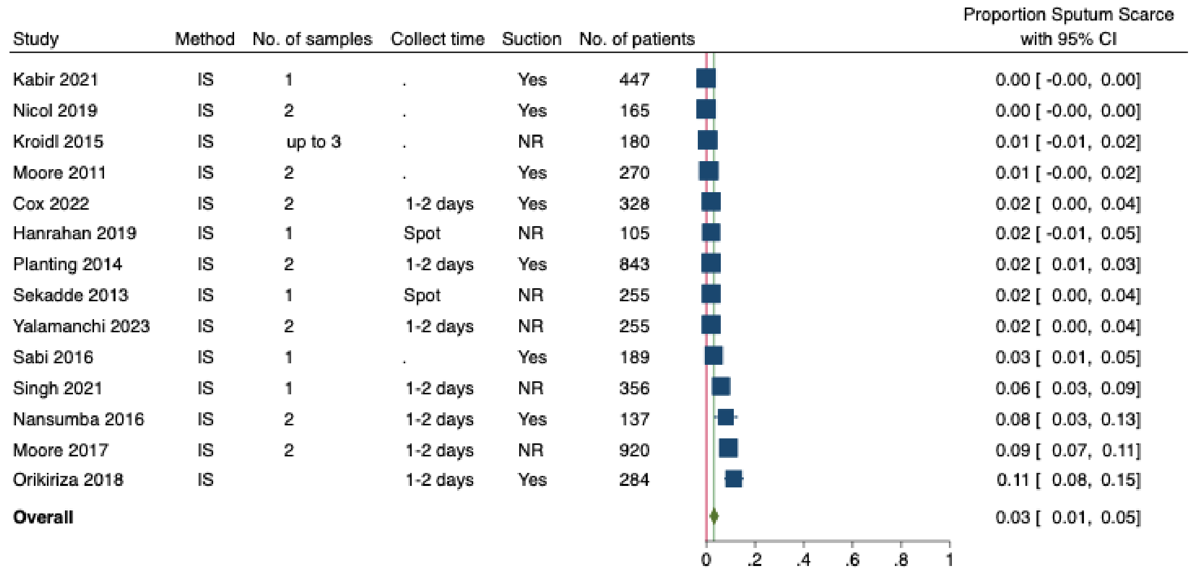

### Figure S7. Meta-analysis of proportion sputum scarcity for collection of induced sputum using suctioning if children are unable to cough, children<15 years

| Study | Method | No. of samples | Collect time | Suction | No. of patients | Proportion Sputum Scarce with 95% CI |
|---|---|---|---|---|---|---|
| Kabir 2021 | IS | 1 | . | Yes | 447 | 0.00 [ -0.00, 0.00] |
| Nicol 2019 | IS | 2 | . | Yes | 165 | 0.00 [ -0.00, 0.00] |
| Moore 2011 | IS | 2 | . | Yes | 270 | 0.01 [ -0.00, 0.02] |
| Cox 2022 | IS | 2 | 1-2 days | Yes | 328 | 0.02 [ 0.00, 0.04] |
| Planting 2014 | IS | 2 | 1-2 days | Yes | 843 | 0.02 [ 0.01, 0.03] |
| Sabi 2016 | IS | 1 | . | Yes | 189 | 0.03 [ 0.01, 0.05] |
| Nansumba 2016 | IS | 2 | 1-2 days | Yes | 137 | 0.08 [ 0.03, 0.13] |
| Orikiriza 2018 | IS | | 1-2 days | Yes | 284 | 0.11 [ 0.08, 0.15] |
| **Overall** | | | | | | 0.03 [ 0.00, 0.06] |

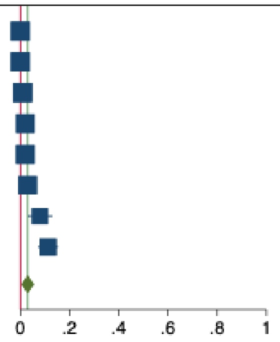
